# A simple laboratory parameter facilitates early identification of COVID-19 patients

**DOI:** 10.1101/2020.02.13.20022830

**Authors:** Qilin Li, Xiuli Ding, Geqing Xia, Zhi Geng, Fenghua Chen, Lin Wang, Zheng Wang

**Author notes:** **Corresponding Authors:** Prof Zheng Wang,; Prof Lin Wang.

## Abstract

The total number of COVID-19 patients since the outbreak of this infection in Wuhan, China has reached 40000 and are still growing. To facilitate triage or identification of the large number of COVID-19 patients from other patients with similar symptoms in designated fever clinics, we set to identify a practical marker that could be conveniently utilized by first-line health-care workers in clinics. To do so, we performed a case-control study by analyzing clinical and laboratory findings between PCR-confirmed SARS-CoV-2 positive patients (n=52) and SARS-CoV-2 negative patients (n=53). The patients in two cohorts all had similar symptoms, mainly fever and respiratory symptoms. The rates of patients with leukocyte counts (normal or decreased number) or lymphopenia (two parameters suggested by current National and WHO COVID-19 guidelines) had no differences between these two cohorts, while the rate of eosinopenia (decreased number of eosinophils) in SARS-CoV-2 positive patients (79%) was much higher than that in SARS-CoV-2 negative patients (36%). When the symptoms were combined with eosinopenia, this combination led to a diagnosis sensitivity and specificity of 79% and 64%, respectively, much higher than 48% and 53% when symptoms were combined with leukocyte counts (normal or decreased number) and/ or lymphopenia. Thus, our analysis reveals that eosinopenia may be a potentially more reliable laboratory predictor for SARS-CoV-2 infection than leukocyte counts and lymphopenia recommended by the current guidelines.

## Introduction

As of Feb 11, 2020, the total number of patients with laboratory-confirmed SARS-CoV-2 infection across China has reached over 40000 and are still growing. ^1^ This outbreak has now spread over 24 countries. The WHO has declared international public health emergency. Referring to previous experience combatting SARS-CoV in 2003 in China, Wuhan and other cities have established fever clinic to triage suspected COVID-19 patients from other patients with similar symptoms. Soon after the outbreak, an overwhelmingly large number of mixed patients with fever or other respiratory symptoms flooded in, resulting in considerably long waiting time for CT examination and etiological PCR tests. Consequently, physicians were often unable to make timely diagnosis for quarantine or therapeutic decisions. A way of accelerating triage process and prioritizing CT examination and PCR tests for suspected SARS-CoV-2 patients would be to identify a useful practical marker. The official guidelines (the Guidelines of the National Health Commission of China for COVID-19, 5^th^ edition;^2^ and the WHO interim guideline^3^) currently recommend two laboratory parameters, “normal/ decreased number of leukocytes” or “decreased number of lymphocytes”, as one of the criteria for diagnosis of COVID-19 infection. Here, our analysis suggests eosinopenia (decreased number of eosinophils) as a potentially more reliable laboratory predictor of SARS-CoV-2 infection than recommended “leukocyte counts” and “lymphopenia”.

## Methods

Data of this retrospective case-negative control study were collected from 105 patients first visiting the Fever Clinic of Wuhan Union Hospital from Feb 3 to Feb 7, 2020. Nasopharyngeal swab specimens of all patients were subject to real time RT-PCR tests through amplifying ORF1ab gene and N gene of SARS-CoV-2 (BioGerm, Shanghai, China). Clinical and laboratory findings were recorded and carefully checked. All statistical analyses were carried out by SPSS 20.0 (SPSS Inc., Chicago, USA). This study was approved by Wuhan Union Hospital Ethics Committee.

## Results

Two cohorts contained SARS-CoV-2-negative (SN) patients (n=53) and SARS-CoV-2-positive (SP) patients (n=52), respectively. The differences in clinical and laboratory findings between the two cohorts were univariately and multivariately analyzed (Table1). No significant differences were found in gender. SP patients (average age 57) was older than SN patients (average age 51, p=0.015). SN patients had a higher rate of other respiratory pathogens infection (34% vs 9.6%, p=0.005). All the patients had pneumonia-like clinical symptoms including fever and respiratory symptoms, whose distribution was similar in the two cohorts. In laboratory findings, the rates of “normal or decreased number of leukocytes” and “lymphopenia” that were used for defining suspected patients by the current guidelines, were not different between these two cohorts. Intriguingly, eosinopenia (<0.02 ×10^9^/ L) was observed in the majority of SP patients (78.8 %), in stark contrast to 35.8 % in SN patients (p<0.001). Consistently, the average eosinophil counts across SP patients (0.02 ×10^9^/ L) was significantly lower than that of SN patients (0.05 ×10^9^/ L, p=0.004).

**Table 1.** Comparison of clinical data and laboratory findings between SARS-CoV-2-negative (SN) and SARS-CoV-2-positive (SP) patients.

|  | SN (n=53) | SP (n=52) | Univariate analysis | Multivariate analysis |  |
| --- | --- | --- | --- | --- | --- |
| **Clinical features** |  |  | **p-value** | **OR** | **p-value** |
| Gender, male/ female | 30/ 23 | 26/ 26 | 0.50 |  | n.s. |
| Age, years | 51 (41-61) | 57 (49-69) | 0.015* |  | n.s. |
| Symptoms of chief complaint |  |  | 0.428 |  |  |
| Fever | 28 (52.8%) | 31 (59.6%) |  |  |  |
| Respiratory symptom | 25 (47.2%) | 20 (38.5%) |  |  |  |
| Weakness | 0 (0) | 1(1.9%) |  |  |  |
| **Laboratory findings** |  |  |  |  |  |
| Other respiratory pathogens infections | 18 (34%) | 5 (9.6%) | 0.005* | 0.05 (0.01 - 0.28) | 0.002* |
| Blood routine examination |  |  |  |  |  |
| Leukocyte (x10<sup>9</sup>/ L; Ref. 3.5 - 9.5) | 5.84 (3.75 - 7.93) | 5.47 (3.74 - 7.2) | 0.32 |  |  |
| Normal or decreased | 49 (92.4%) | 50 (96.2%) | 0.33 |  |  |
| Neutrophil (x10<sup>9</sup>/ L; Ref. 1.8 - 6.3) | 3.98 (2.39 - 5.57) | 3.74 (2.18 -5.3) | 0.49 |  |  |
| Increased | 5 (9.4%) | 3 (5.7%) | 0.65 |  |  |
| Lymphocyte (x10<sup>9</sup>/ L; Ref. 1.1 - 3.2) | 1.38 (0.66 - 2.09) | 1.23 (0.68 - 1.78) | 0.11 |  |  |
| Decreased | 25 (47.2%) | 25 (48.1%) | 0.93 |  |  |
| Monocytes (x10<sup>9</sup>/ L; Ref. 0.1 - 0.6) | 0.43 (0.21 - 0.64) | 0.45 (0.28 -0.68) | 0.68 |  |  |
| Increased | 8 (15.1%) | 12 (23.1%) | 0.30 |  |  |
| Eosinophil (x10<sup>9</sup>/ L; Ref. 0.02 - 0.52) | 0.05 (-0.04 - 0.14) | 0.02 (-0.01 - 0.05) | 0.004* |  |  |
| Decreased | 19 (35.8%) | 41 (78.8%) | <0.001* | 0.10 (0.03 - 0.32) | < 0.001* |
| Red blood cell count (x10<sup>12</sup>/ L; Ref. 3.8 - 5.1) | 4.72 (4.21 - 5.23) | 4.71 (4.25 - 5.12) | 0.93 |  |  |
| Hemoglobin (g/ L; Ref. 115 -150) | 140.8 (125.1 - 156.5) | 140.8 (127.3 -154.3) | 0.99 |  | n.s. |
| Platelet (x10<sup>9</sup>/ L; Ref. 125 - 350) | 237.5 (135.2 - 335.9) | 198.4 (125.8-270.9) | 0.022* |  | n.s. |
| hCRP (mg/ L; Ref. < 4) | 23.6 (-6.9 - 54.1) | 36.5 (-2.8 - 75.8) | 0.32 |  | n.s. |
| Increased | 36 (67.9%) | 42 (80.8%) | 0.14 |  |  |

Data are presented as n/ n, n (%), media (IQR) and mean (95% CI). Other respiratory pathogen infections include mycoplasma, chlamydia, syncytial virus, adenovirus and coxsackie virus B, and influenza. Ref., normal reference range; OR, odds ratio; hCRP, hypersensitive C-reactive protein; \*, p - value < 0.05, significantly different.

The performance of eosinopenia as a predictor for SARS-CoV-2 infection was then determined in the presence of symptoms (Table 2). The inclusion of leukocyte counts (normal or decreased number) had no significant diagnostic value. Notably, the inclusion of eosinopenia remarkably improved the sensitivity and specificity to 78.8% and 64.2%, respectively, much higher than the inclusion of lymphopenia (48.1% and 52.8%). Further, the combination of other blood parameters (leukocytes and/ or lymphopenia) with eosinopenia increased selectivity but reduced sensitivity.

**Table 2.** Diagnostic performance of single and combined laboratory parameters in conjunction with symptoms on differentiating SARS-CoV-2-positive patients from SARS-CoV-2-negative patients.

|  | **Sensitivity (%)** | **Specificity (%)** | **PPV (%)** | **NPV (%)** |
| --- | --- | --- | --- | --- |
| Symptoms + Leukocytes ($\leq 3.5$) | 96.2 | 5.7 | 50.0 | 60.0 |
| Symptoms + lymphopenia ($< 1.1$) | 48.1 | 52.8 | 50.0 | 50.9 |
| Symptoms + lymphopenia + Leukocytes ($\leq 3.5$) | 46.2 | 52.8 | 49.0 | 50.0 |
| Symptoms + eosinopenia ($< 0.02$) | 78.8 | 64.2 | 68.3 | 75.6 |
| Symptoms + eosinopenia ($< 0.02$) + leukocytes ($\leq 3.5$) | 75.0 | 66.0 | 68.4 | 72.9 |
| Symptoms + eosinopenia ($< 0.02$) + lymphopenia ($< 1.1$) | 38.5 | 75.5 | 60.6 | 55.6 |
| Symptoms + eosinopenia ($< 0.02$) + lymphopenia ($< 1.1$) + leukocyte ($\leq 3.5$) | 36.5 | 75.5 | 59.4 | 54.8 |

Numbers are absolute cell counts ( $10^9/\text{L}$ ). PPV, positive predictive value; NPV, negative predictive value.

## Discussion

Eosinopenia can be observed in typhoid fever or as a response to glucocorticoid. In previous clinical reports on SARS-CoV, MERS-CoV and current SAR-CoV-2,^4-6^ changes in eosinophils in peripheral blood were usually omitted. Our study showed that eosinopenia appeared in the majority of SP patients. The mechanisms underlying such eosinopenia were currently unclear and warrant further study.

Since the data was collected at patients’ first medical visit, eosinopenia was presumed to be an early event in patients’ clinical course, possibly prior to emergence of characteristic radiological findings. Combined with fever and respiratory symptoms, eosinopenia as a parameter in routine blood tests might be capable of facilitating rapid identification of highly-suspected cases from mixed patients at triage in fever clinics. These quickly-identified suspected patients would be recommended to (1) receive the priority for radio-diagnosis and laboratory-definitive diagnosis, (2) be isolated in designated wards without any delay to avoid potential virus spreading, (3) and receive empirical antiviral treatment (as of now no consensus over usage of antiviral agents) to prevent aggravation. The additional inclusion of eosinopenia may further refine the laboratory diagnostic criteria recommended by the current guidelines.

## Data Availability

All the data referred to in this manuscript are available upon appropriate requests.

## Author Contributions

Dr Q. Li, X. Ding, and Prof. G. Xia contributed equally to this work. All corresponding authors had full access to all the data in the study and take responsibility for the integrity of the data and the accuracy of the data analysis.

*Study concept and design:* Z. Wang, L. Wang, G. Xia

*Acquisition, analysis, or interpretation of data:* All authors.

*Drafting of the manuscript:* Z. Wang, L. Wang, Q. Li, X. Ding

*Critical revision of the manuscript for important intellectual content:* Z. Geng, F. Chen. G. Xia

*Supervision:* Z. Wang, L. Wang.

